# Proxalutamide Improves Lung Injury in Hospitalized COVID-19 Patients – an Analysis of the Radiological Findings of the Proxa-Rescue AndroCoV Trial

**DOI:** 10.1101/2021.07.01.21259656

**Authors:** Flávio Adsuara Cadegiani, Daniel do Nascimento Fonseca, Michael do Nascimento Correia, Renan Nascimento Barros, Dirce Costa Onety, Karla Cristina Petruccelli Israel, Emilyn Oliveira Guerreiro, José Erique Miranda Medeiros, Raquel Neves Nicolau, Luiza Fernanda Mendonça Nicolau, Rafael Xavier Cunha, Maria Fernanda Rodrigues Barroco, Patrícia Souza da Silva, Raysa Wanzeller de Souza Paulain, Claudia Elizabeth Thompson, Ricardo Ariel Zimerman, Carlos Gustavo Wambier, Andy Goren

**Affiliations:** Corpometria Institute, Brasilia, Brazil; Applied Biology, Inc. Irvine, CA, USA; Samel & Oscar Nicolau Hospitals, Manaus, Brazil; Hospital Regional José Mendes, Itacoatiara, Amazonas, Brazil; Hospital Municipal Jofre Cohen, Parintins, Amazonas, Brazil; Centro de Doenças Renais do Amazonas, Manaus, Brazil; Programa de Pós-Graduação em Medicina Tropical – FMT/UEA, Manaus, Brazil; Department of Pharmacosciences, Universidade Federal de Ciências da Saúde de Porto Alegre; Hospital da Brigada Militar, Porto Alegre, Brazil; Department of Dermatology, Alpert Medical School of Brown University, Providence, RI, USA

**Keywords:** COVID-19, SARS-CoV-2, proxalutamide, antiandrogen, non-steroidal antiandrogen (NSAA), lung injury

## Abstract

**Introduction:** Antiandrogens are candidates against the severe acute respiratory syndrome coronavirus 2 (SARS-CoV-2) disease (COVID-19) due to host cell entry inhibition by the suppression of TMPRSS2. Proxalutamide is a nonsteroidal anti-androgen (NSAA) with strong antagonism on androgen receptor (AR) and angiotensin-converting enzyme 2 (ACE2). Efficacy of proxalutamide was previously demonstrated for early COVID-19 outpatients, and also reduction of deaths in hospitalized COVID-19 patients. Whether radiological changes would follow the improvement in clinical outcomes with proxalutamide is not established. The present *post-hoc* analysis aims to evaluate whether proxalutamide improves lung injury observed through chest computerized tomography (CT) scans.

**Methods:** This is a *post-hoc* analysis of the radiological findings of The Proxa-Rescue AndroCoV Trial with all enrolled patients from the three participating institutions of the city of Manaus, Brazil, that had at least two chest CT scans during hospitalization. The quantification of lung parenchyma involvement was performed by blinded radiologists with expertise in analysis of COVID-19 images. A first chest CT scan was performed upon randomization and a second CT scan was performed approximately five days later, whenever feasible. Improvement rate was the first endpoint, and relative and absolute changes between the first and second CT scans were the second endpoints.

**Results:** Of the 395 patients initially evaluated, 77 and 169 patients from the proxalutamide and placebo arms, respectively, were included (n=246). Baseline characteristics and percentage of lung parenchyma affected in the baseline chest CT scan were similar between groups. In the second chest CT scan, the percentage of lungs affected (Median – IQR) was 35.0% (25.0-57.5%) in the proxalutamide group versus 67.5% (50.0-80.0%) in the placebo group (p < 0.001). The absolute and relative change between the second and first chest CT scans (Median – IQR) were -15.0 percent points (p.p.) (−30.0 – 0.0p.p.) and -25.0% (−50.0 – 0.0%) in the proxalutamide group, respectively, and +15.0p.p. (0.0 - +30.0p.p.) and +32.7% (0.0 - +80.0%) in the placebo group, respectively (p < 0.001 for both absolute and relative changes). The improvement rate, *i*.*e*., the percentage of subjects that had improvement from the first to the second CT scan, was 72.3% in the proxalutamide group and 23.1% in the placebo group (p < 0.0001), with an improvement rate ratio (95%CI) of 3.15 (2.32 – 4.28).

**Conclusion:** Proxalutamide improves lung opacities in hospitalized COVID-19 patients when compared to placebo. (NCT04728802)

## Introduction

Antiandrogen drugs are therapeutic candidates against the severe acute respiratory syndrome coronavirus 2 (SARS-CoV-2) disease (COVID-19) due to inhibition of cell-entry mechanisms.^1^ The blockade of viral cell entry would occur indirectly by the mitigation of the structural modification of the SARS-CoV-2 spike (S) protein that is required for infection. This inhibition occurs though the reduction of the expression of the endogenous transmembrane protease, serine 2 (TMPRSS2). The enzyme is responsible for priming the viral S protein, allowing its proper coupling to angiotensin-converting enzyme 2 (ACE2) receptor and consequent cell entry.^2^ The suppression of the TMPRSS2 expression occurs through the inhibition of the TMPRSS2 promoter, that includes an androgen response element.^3^ Since androgens are the only known endogenous regulators of TMPRSS2, antiandrogens play a key role in the inhibition of TMPRSS2 expression. Pre-clinical studies have shown that nonsteroidal antiandrogens down regulate TMPRSS2^4^ and inhibit viral replication in human cell culture.^5,6^ However, other mechanisms to explain the potential interplay between antiandrogens and SARS-CoV-2, such as *downregulation* of ACE2 receptors, may play an additional role, and should be further elucidated.

Proxalutamide is a second-generation nonsteroidal anti-androgen (NSAA) that is more potent than other NSAAs, such as enzalutamide or bicalutamide.^7^ In addition to the competitive antagonism in the androgen receptor (AR), NSAAs also prevent androgen receptor nuclear translocation and binding to DNA.^8^

The efficacy of proxalutamide was previously demonstrated for SARS-CoV-2 positive men in an outpatient setting.^9,10^ The Proxa-Rescue AndroCoV Trial has demonstrated the efficacy of proxalutamide for hospitalized COVID-19 men and women, with 128% increase in 14-day alive hospital discharge, and 77.7% reduction in the 28-day mortality rate, though a double-blinded, placebo-controlled, multicenter randomized clinical trial.^11^

Radiological improvement of COVID-19 tends to occur later in the recovery process, with easing of lungs appearance on chest computed tomography (CT) scan usually observed only after seven to 14 days.^12^ Due to the fast improvement observed in patients of the proxalutamide arm, we hypothesized that an earlier improvement in the radiological aspect on the chest CT scan could also occur in hospitalized COVID-19 patients.

The objective of the present study is to compare the CT scans between the proxalutamide and placebo arms, through blinded radiologists’ evaluation and reading of CT scans during the course of the RCT of hospitalized COVID-19 patients.

## Methods

### Trial Design, Setting and Locations

Study design, criteria for eligibility, randomization, procedures, outcomes are described elsewhere.^11^

This is a *post-hoc* analysis of the radiological findings of a double-blinded, placebo-controlled, prospective, two-arm randomized clinical trial (RCT), that encompassed the three institutions of the city of Manaus, Amazonas, Brazil, that participated in the study. The other five centers included in the RCT were not included due to the lack of available CTs for regular evaluation of COVID-19. The study was conducted between February 1 and April 15, 2021, including enrollment and follow-up.

The RCT was approved by Brazilian National Ethics Committee of the Ministry of Health, under the approval number 4.513.425 of the process number (CAAE) 41909121.0.0000.5553 (original name of the Ethics Committee: Comitê de Ética em Pesquisa (CEP) do the Comitê Nacional de Ética em Pesquisa (CONEP) do Ministério da Saúde - CEP/CONEP/MS). All data used for the present *post-hoc* analysis was entirely covered by the approval obtained with the Brazilian National Ethics Committee of the Ministry of Health (MS) (approval number 4.513.425). The RCT was registered in clinicaltrials.gov (NCT04728802).

### Eligibility criteria

In short, for inclusion, men and women above 18 years old hospitalized due to COVID-19 confirmed with a positive real-time reverse transcription polymerase chain reaction (rtPCR) test for SARS-CoV-2 (Cobas SARS-CoV-2 rtPCR kit test protocol, Roche, USA) were considered.

Exclusion criteria included mechanical ventilation at the time of randomization, known congestive heart failure class III or IV (New York Heart Association), immunosuppression, alanine transferase (ALT) above five times ULN (> 250 U/L), creatinine above 2.5 mg/ml or a calculated eGFR below 30 ml/min, current use of antiandrogen medications, planning to attempt to have kids within 90 days after the intervention, and women that were pregnant or breastfeeding.

For the present *post-hoc* analysis of the radiological findings, all patients that participated in the Proxa-Rescue AndroCoV Trial^11^ from hospitals with CT available were included, which include the three hospitals located in the city of Manaus, Brazil, since none of the other hospitals had CTs in their facilities. Patients with at least two chest CT scans with known percentage of affected lung parenchyma during hospitalization were included in the present analysis, since at least two scans were needed for comparison purposes. All potential limitations of a subgroup *post-hoc* analysis of a RCT described by Pocock *et al* were addressed.^13^

### Procedures

Patients were randomized to receive either proxalutamide 300 mg/day plus usual care or a placebo plus usual care for 14 days in a 1:1 ratio. If patients were discharged before 14 days, they were instructed to continue treatment. Therapy compliance was monitored daily for both inpatients and patients that were discharged until day 14, and then in days 21 and 28 if discharged before, or daily if still hospitalized.

The COVID-19 8-point ordinal scale was used as the parameter for monitoring. The ordinary clinical scale is defined as: 8. Death; 7. Hospitalized, on invasive mechanical ventilation; 6. Hospitalized, on non-invasive ventilation or high flow oxygen devices; 5. Hospitalized, requiring supplemental oxygen; 4. Hospitalized, not requiring supplemental oxygen-requiring ongoing medical care (COVID-19 related or otherwise); 3. Hospitalized, not requiring supplemental oxygen - no longer requires ongoing medical care; 2. Not hospitalized, limitation on activities; and 1. Not hospitalized, no limitations on activities.^11^

Baseline characteristics, previous medical history, comorbidities and concomitant medications were recorded. Usual care for hospitalized COVID-19 patients as per the hospitals protocol included enoxaparin, colchicine, methylprednisolone or dexamethasone, and antibiotic therapy as required. The usual care was not changed for the RCT.

Before the onset of the RCT, a random sequence using 4, 6 and 8 block sizes and a list length for 662 treatments was created thought a randomization software.^14^ The randomization sequence and allocation concealment were performed remotely and was not stratified by institution. Pre-packing of tablets of either active or placebo group was manufactured to have identical physical characteristics, and was manufactured and transported by Kintor Pharmaceuticals Ltd. Suzhou, China.

### Protocol for the exploratory analysis of the chest CT scans

As per the protocol of the three hospitals located in the city of Manaus, Amazonas, Brazil, patients hospitalized due to COVID-19 had chest CT-scans approximately every five days, or whenever it was feasible to transport patient to the CT-scan room. A first chest CT scan was performed upon randomization and a second CT scan was performed approximately five days later, whenever patient health condition permitted the transportation. Patients in ICU or clinically unstable were not eligible for the five-day interval CT-scan follow-up.

The analysis of the chest CT scan was performed by board-certified radiologists with previous clinical expertise in COVID-19, that quantified the percentage of lung parenchyma involved in COVID-19, based on the classifications proposed by Xie et al, Zhao et al, Pan et al, Li et al, Chung et al, and Yuan et al,^15-20^ following the standardization proposed by Martinez Chamorro et al.^12^ The three hospitals unified and standardized the methods for the quantification of lung affected in order to avoid interoperator differences. All chest CT scans were performed in CT SOMATOM model with 64-slice data acquisition (Simens Healthineers, Siemens, Germany).

Bilateral reticular patterns, peripheral bilateral ground-glass opacities, and patchy or confluent multifocal consolidation were considered as findings consistent with COVID-19 pneumonia. Central consolidation and unilateral ground-glass opacities were considered as indeterminate for COVID-19. Pneumothorax, pneumomediastinum, pleural effusion, lobar consolidation, military patterns or cavitation were not considered as part of COVID-19 pneumonia, although a series of case reports have described the first two characteristics. Long term fibrotic changes, such as honeycombing or traction bronchiectasis, could be consequences of COVID-19, but were not considered as part of the quantification of lungs affected. In short, only CO-RADS 6 were included in this analysis. All patients had diagnosis of COVID-19 through positive rtPCR-SARS-CoV-2 test.^21^

The analyses were performed in a complete independent manner. Radiologists were not informed whether patients were or were not participating in the RCT, thus, were also blinded regarding the allocation in the proxalutamide or placebo arms.

For quantification purposes, whenever an interval of percentage was provided instead of an exact percentage, this was replaced by an exact value, as following: <5% = 2%; <10% = 5%: <25% = 10%; 10-25% = 20%; <30% = 15%; <50% = 30%; 25-50% = 40%; >30% = 50%: 50-60% = 60%; >50% = 70%; 50-75% = 65%; >75% = 90%; >80% = 90%; and >90% = 95%.

For the present exploratory analysis, all 395 COVID-19 hospitalized patients enrolled in the RCT from the three hospitals of the city of Manaus, Amazonas, were initially considered, without any sort of previous selection, to avoid selection bias. Among these patients, all those with at least two chest CT scans during hospitalization with known results were included for the analysis.

### Endpoints

Primary endpoint was improvement rate (percentage of subjects that presented improvement from the first to the second chest CT scan) of each group, and the measure of efficacy used was Improvement Ratio (IR). Second endpoints included differences in the quantification of lung parenchyma affected by COVID-19 chest, seen through chest CT scan results, between the baseline (first) and second exam, in terms of: 1. absolute changes (in points percent – pp; *eg*.. If the first CT scan showed 50% of lungs affected and the second CT scan showed 25% of lungs affected, a 25pp reduction – -25pp – was observed); and 2. relative changes (percentage of change.; *eg*.. If the first CT scan showed 50% of lungs affected and the second CT scan showed 25% of lungs affected, a 50%. reduction – -50% – was observed, compared to the first CT scan), were compared between proxalutamide and placebo arms; and 14-day and 28-day all-cause mortality rate of each group.

Baseline characteristics and clinical outcomes are described to evaluate whether the dimension of the drug efficacy of the RCT is represented in this *post-hoc* analysis.

### Availability of dataset

For reproduction purposes, we provide the unidentified dataset in the Supplement 1.

### Statistical Analysis

An original intention-to-treat (ITT) protocol (unmodified) was used for data analysis. Since this is a post-hoc analysis of the subgroup of patients who were in hospitals with CT scans available, a post-hoc power calculation was performed for the primary findings. With the sample size of the present analysis, an improvement of at least 2.0 in terms of improvement rate ratio would lead to a statistical power of 95%.

Analysis was also stratified by sex in order to detect whether improvements would be sex-specific or not, although males and females presented similar clinical responses to proxalutamide in the RCT, when compared to placebo.

Cox proportional hazards model was used to calculate hazard ratio (HR) for all-cause 14-day and 28-day mortality and their 95% confidence interval (CI), to measure the effects of proxalutamide versus placebo. To measure the effects of proxalutamide *versus* placebo in terms of radiological endpoints, normality tests (skewness and kurtosis tests) were performed for all variables to determine with statistical tools would be employed. When distribution was normal, parametric ANOVA t-test was performed, and data was provided as mean and 95% confidence interval (95%CI). When data was not normally distributed, non-parametric Kruskal-Wallis Test was performed and data was provided in median and interquartile range (IQR). Differences were evaluated and disclosed as *p-values*, and statistical significance was established at p < 0.05. Improvement rate ratios were calculated using Cox proportional hazards model. All statistical tests were performed using IBM-SPSS statistics version 25.0 software (IBM, USA).

## Results

The flowchart depicting the subject selection in the proposed protocol is shown in Figure 1. A total of 395 patients were initially evaluated, including 97 patients in the active arm and 298 patients in the placebo arm. Of these, 77 (79.4%) of the proxalutamide group and 169 (56.7%) of the placebo group had at least two chest CT scans with known percentage of affected lung parenchyma (p = 0.0006). The median interval between two chest CT scans was 5.0 days for both groups (p = n/s). Imbalances between sites in terms of active and placebo proportions are explained elsewhere.^11^

**Figure 1.**
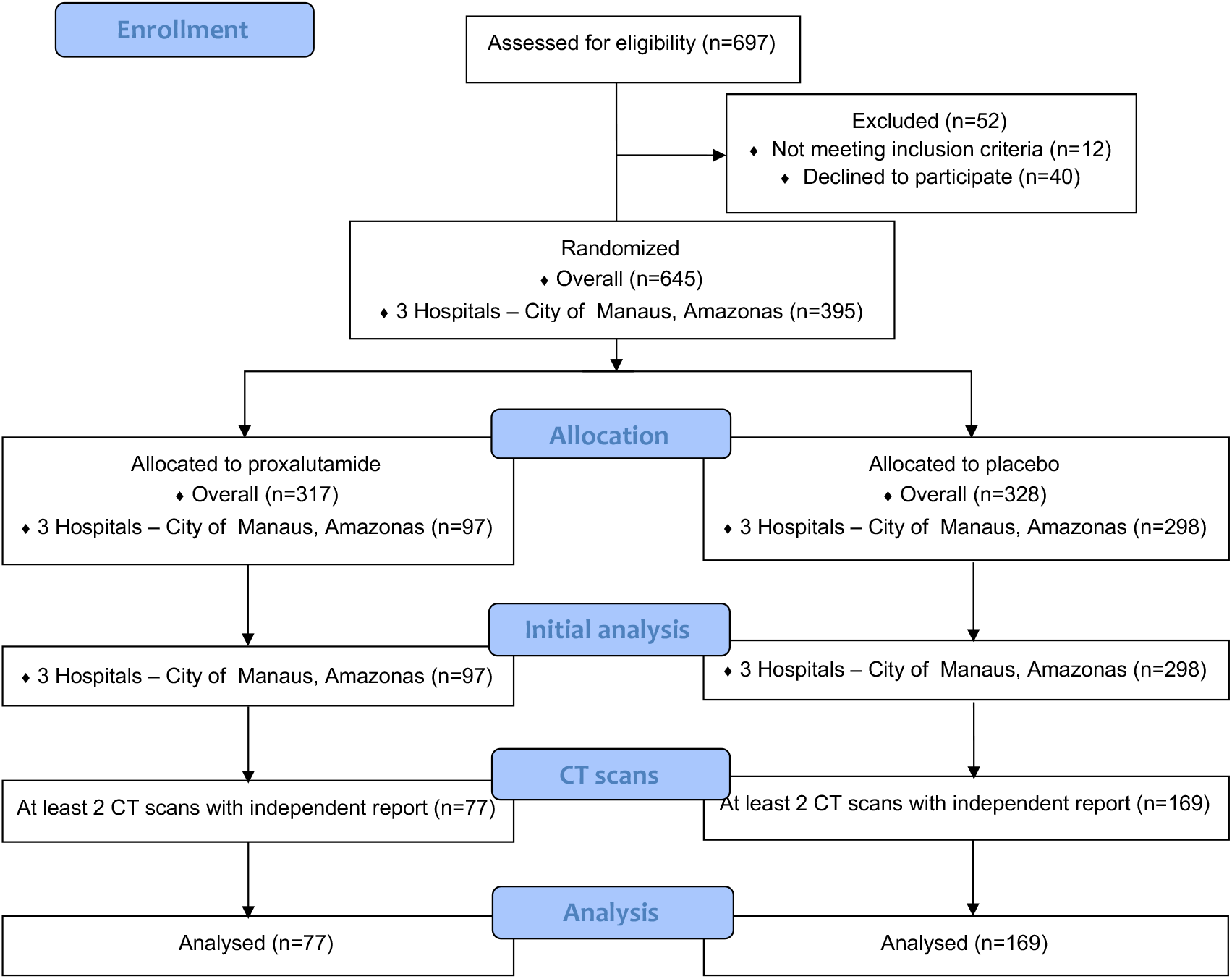
Procotol flowchart – Analysis of the chest computerized tomography scans in the Proxa-Rescue AndroCoV Trial.

Baseline characteristics, including age, proportion between males and females, and presence of comorbidities, and additional parameters such as median time since hospitalization, distribution of the score in the COVID-19 ordinary scale, and use of concomitant medications were similar between proxalutamide and placebo group (Table 2).

**Table 1.** Selection of patients

CT = computerized tomography; IQR = interquartile range
| Parameter | Overall<br>N = 395 | Proxalutamide<br>N = 97 | Placebo<br>N = 298 | p |
| --- | --- | --- | --- | --- |
| *Female* | 170 (43.0%) | 38 (39.2%) | 132 (44.3%) | 0.41 |
| *Male* | 225 (57.0%) | 59 (60.8%) | 166 (55.7%) |  |
| Number of subjects with at least 1 Chest CT scan – n (%) | 357 (90.4%) | 89 (91.7%) | 268 (89.9%) | 0.69 |
| *Female* | 148 (88.1%) | 35 (92.1%) | 113 (85.6%) | 0.41 |
| *Male* | 209 (92.1%) | 54 (91.5%) | 155 (93.4%) | 0.77 |
| Number of subjects with at least 2 Chest CT scans – n (%) | 250 (63.3%) | 78 (80.4%) | 172 (57.7%) | **0.0001** |
| *Female* | 96 (57.1%) | 31 (81.6%) | 65 (49.2%) | **0.0004** |
| *Male* | 154 (67.8%) | 47 (79.7%) | 107 (64.5%) | **0.034** |
| Number of subjects with at least 2 Chest CT scans with known % of lung parenchyma affected – n (%) | 246 (62.3%) | 77 (79.4%) | 169 (56.7%) | **0.0001** |
| *Female* | 96 (57.1%) | 31 (81.6%) | 65 (49.2%) | **0.0004** |
| *Male* | 154 (67.8%) | 46 (78.0%) | 104 (62.6%) | **0.037** |
| Interval (days) between first and second chest CT scan - Median (IQR) | 5.0<br>(4.0-7.0) | 5.0<br>(4.0 – 6.0) | 5.0<br>(4.0 – 7.0) | 0.13 |
| *Female* | 5.0<br>(4.0 – 7.0) | 5.0<br>(4.0 – 6.0) | 5.0<br>(4.0 – 7.0) | 0.14 |
| *Male* | 5.0<br>(4.0 – 7.0) | 5.0<br>(4.0 – 6.0) | 5.0<br>(4.0 – 8.0) | 0.53 |

**Table 2.**
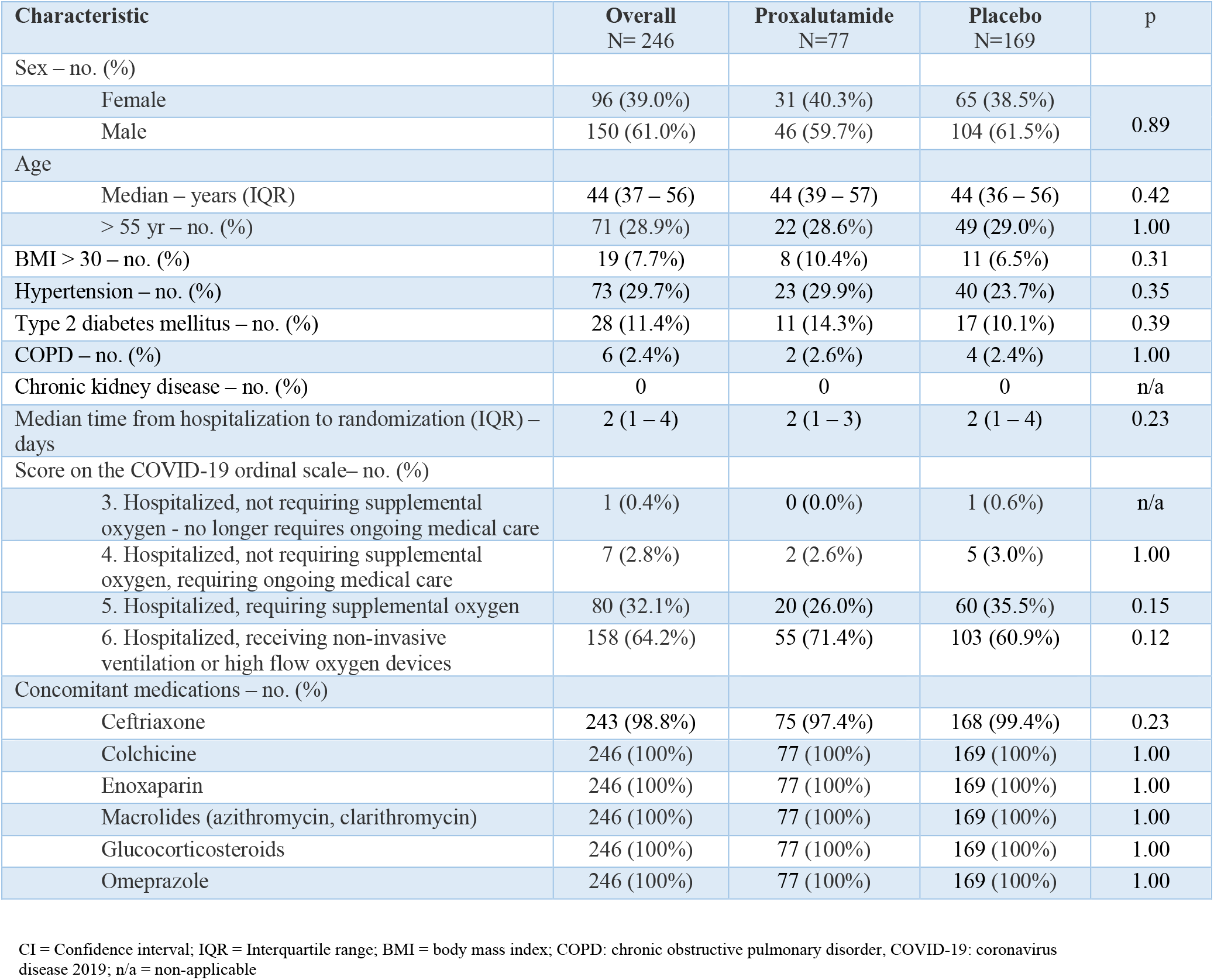
Baseline clinical characteristics, outcomes, and adverse effects.

| Characteristic | Overall<br>N= 246 | Proxalutamide<br>N=77 | Placebo<br>N=169 | p |
| --- | --- | --- | --- | --- |
| Sex – no. (%) |  |  |  |  |
| Female | 96 (39.0%) | 31 (40.3%) | 65 (38.5%) | 0.89 |
| Male | 150 (61.0%) | 46 (59.7%) | 104 (61.5%) |  |
| Age |  |  |  |  |
| Median – years (IQR) | 44 (37 – 56) | 44 (39 – 57) | 44 (36 – 56) | 0.42 |
| > 55 yr – no. (%) | 71 (28.9%) | 22 (28.6%) | 49 (29.0%) | 1.00 |
| BMI > 30 – no. (%) | 19 (7.7%) | 8 (10.4%) | 11 (6.5%) | 0.31 |
| Hypertension – no. (%) | 73 (29.7%) | 23 (29.9%) | 40 (23.7%) | 0.35 |
| Type 2 diabetes mellitus – no. (%) | 28 (11.4%) | 11 (14.3%) | 17 (10.1%) | 0.39 |
| COPD – no. (%) | 6 (2.4%) | 2 (2.6%) | 4 (2.4%) | 1.00 |
| Chronic kidney disease – no. (%) | 0 | 0 | 0 | n/a |
| Median time from hospitalization to randomization (IQR) – days | 2 (1 – 4) | 2 (1 – 3) | 2 (1 – 4) | 0.23 |
| Score on the COVID-19 ordinal scale– no. (%) |  |  |  |  |
| 3. Hospitalized, not requiring supplemental oxygen - no longer requires ongoing medical care | 1 (0.4%) | 0 (0.0%) | 1 (0.6%) | n/a |
| 4. Hospitalized, not requiring supplemental oxygen, requiring ongoing medical care | 7 (2.8%) | 2 (2.6%) | 5 (3.0%) | 1.00 |
| 5. Hospitalized, requiring supplemental oxygen | 80 (32.1%) | 20 (26.0%) | 60 (35.5%) | 0.15 |
| 6. Hospitalized, receiving non-invasive ventilation or high flow oxygen devices | 158 (64.2%) | 55 (71.4%) | 103 (60.9%) | 0.12 |
| Concomitant medications – no. (%) |  |  |  |  |
| Ceftriaxone | 243 (98.8%) | 75 (97.4%) | 168 (99.4%) | 0.23 |
| Colchicine | 246 (100%) | 77 (100%) | 169 (100%) | 1.00 |
| Enoxaparin | 246 (100%) | 77 (100%) | 169 (100%) | 1.00 |
| Macrolides (azithromycin, clarithromycin) | 246 (100%) | 77 (100%) | 169 (100%) | 1.00 |
| Glucocorticosteroids | 246 (100%) | 77 (100%) | 169 (100%) | 1.00 |
| Omeprazole | 246 (100%) | 77 (100%) | 169 (100%) | 1.00 |

CI = Confidence interval; IQR = Interquartile range; BMI = body mass index; COPD: chronic obstructive pulmonary disorder, COVID-19: coronavirus disease 2019; n/a = non-applicable

Table 3 describes the outcomes and adverse effects for the population analyzed. Of the patients included in the present exploratory analysis, the 14-day mortality was 1.3% in the proxalutamide group and 21.3% in the placebo group, with a mortality risk ratio (RR) of 0.06 (0.01-0.43). The 28-day mortality was 2.6% in the proxalutamide group and 31.4% in the placebo group, with a mortality RR of 0.08 (0.02-0.33). The median (interquartile range – IQR) hospitalization length stay was eight (6 – 10) days in the proxalutamide group and 14 (9 – 19) days in the placebo group (p < 0.0001). The only adverse effect more commonly present in the proxalutamide group was diarrhea (20.8% versus 3.6%, p = 0.008).

**Table 3.** Outcomes and adverse effects of the population analyzed.

| Parameter | Overall<br>N= 246 | Proxalutamide<br>N=77 | Placebo<br>N=169 | p |
| --- | --- | --- | --- | --- |
| 14-day all-cause mortality – no. (%) | 37 (15.0%) | 1 (1.3%) | 36 (21.3%) | < 0.0001 |
| 14-day all cause mortality rate ratio | 0.06 (0.01 – 0.43) |  |  |  |
| 28-day all-cause mortality – no. (%) | 55 (22.4%) | 2 (2.6%) | 53 (31.4%) | < 0.0001 |
| 28-day all cause mortality rate ratio | 0.08 (0.02 – 0.33) |  |  |  |
| Score on the COVID-19 ordinal scale at Day 14– Median (IQR) | 2 (1 – 5) | 1 (1 – 1) | 3 (2 – 7) | < 0.0001 |
| Score on the COVID-19 ordinal scale at Day 28– Median (IQR) | 1 (1 – 3) | 1 (1 – 1) | 2 (1 – 8) | < 0.0001 |
| Median hospitalization days (IQR) | 11 (7 – 17) | 8 (6 – 10) | 14 (9 – 19) | < 0.0001 |
| Median hospitalization days after randomization (IQR) | 8 (5 – 14) | 6 (4 – 8) | 10 (6 – 16) | < 0.0001 |
| **Grade 5 – n (%)** |  |  |  |  |
| Death, Day 14 | 37 (15.0%) | 1 (1.3%) | 36 (21.3%) | < 0.0001 |
| Death, Day 28 | 55 (22.4%) | 2 (2.6%) | 53 (31.4%) | < 0.0001 |
| **Grades 4 or 3 – n (%)** |  |  |  |  |
| Mechanical ventilation, Day 14 | 20 (8.1%) | 0 | 20 (11.8%) | < 0.0001 |
| Mechanical ventilation, Day 28 | 1 (0.4%) | 0 | 1 (0.6%) | n/a |
| Renal failure (creatinine increase > 100%) | 8 (3.3%) | 2 (2.6%) | 6 (3.6%) | 1.00 |
| Liver damage (ALT > 250 U/L or >100% increase) | 8 (3.3%) | 2 (2.6%) | 6 (3.6%) | 1.00 |
| **Grades 2 or 1 – n (%)** |  |  |  |  |
| Diarrhea | 22 (8.9%) | 16 (20.8%) | 6 (3.6%) | 0.008 |
| Abdominal pain | 2 (0.8%) | 1 (1.3%) | 1 (0.6%) | n/a |
| Irritability | 1 (0.4%) | 1 (1.3%) | 0 (0.0%) | n/a |
| Spontaneous erection | 1 (0.4%) | 1 (1.3%) | 0 (0.0%) | n/a |
| Vomiting, dyspepsia, or palpitations | 0 | 0 | 0 | - |

IQR = Interquartile range; n/a = non-applicable

Radiological findings of overall subjects and for each group are described in Table 4. Figure 2 and Figure 3 illustrate the changes in CT scans in both groups, and baseline and during-treatment CT scans in proxalutamide versus placebo group, respectively.

**Table 4.** Radiological outcomes.

| Parameter | Overall<br>N = 246 | Proxalutamide<br>N = 77* | Placebo<br>N = 169* | p |
| --- | --- | --- | --- | --- |
| **First** CT scan (% of affected lungs) - Median (IQR) | 50<br>(40 – 70) | 60<br>(43 – 70) | 50<br>(30 – 70) | n/s |
| **Second** CT scan (% of affected lungs) - Median (IQR) | 55 (40 – 70) | 35 (25 – 60) | 68 (50 – 80) | < 0.0001 |
| **Improvement rate - % (n)** | 38.6% (95) | 72.3% (56) | 23.1% (39) | < 0.0001 |
| **Improvement rate ratio (95% CI)** | 3.15 (2.32 – 4.28) |  |  |  |
| **Absolute change (pp)** between first and second chest CT scan - Median (IQR) | 0pp<br>(-10pp – +20pp) | -15pp<br>(-30pp – 0pp) | +15pp<br>(0 pp - +30pp) | < 0.001 |
| **Relative change (%)** in terms of percentage between first and second chest CT scan - Median (IQR) | 0.0%<br>(-30% – +20%) | -25%<br>(-50% – 0%) | +33%<br>(0% - +80%) | < 0.001 |

CT = computerized tomography; IQR = interquartile range; pp = points percentage
\* Included for analysis;

**Figure 2.**
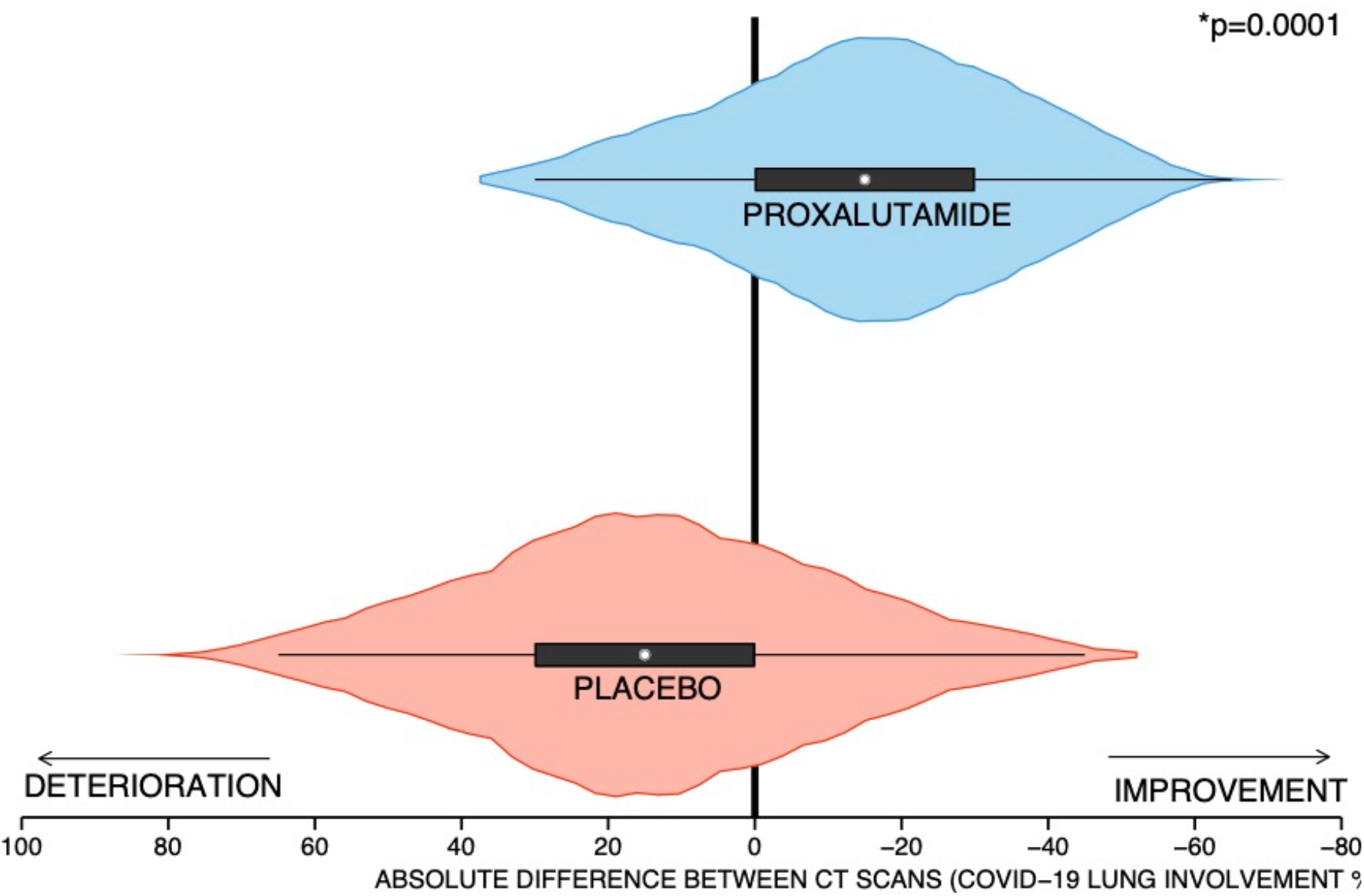
Changes between the first and second CT scans.

**Figure 3.**
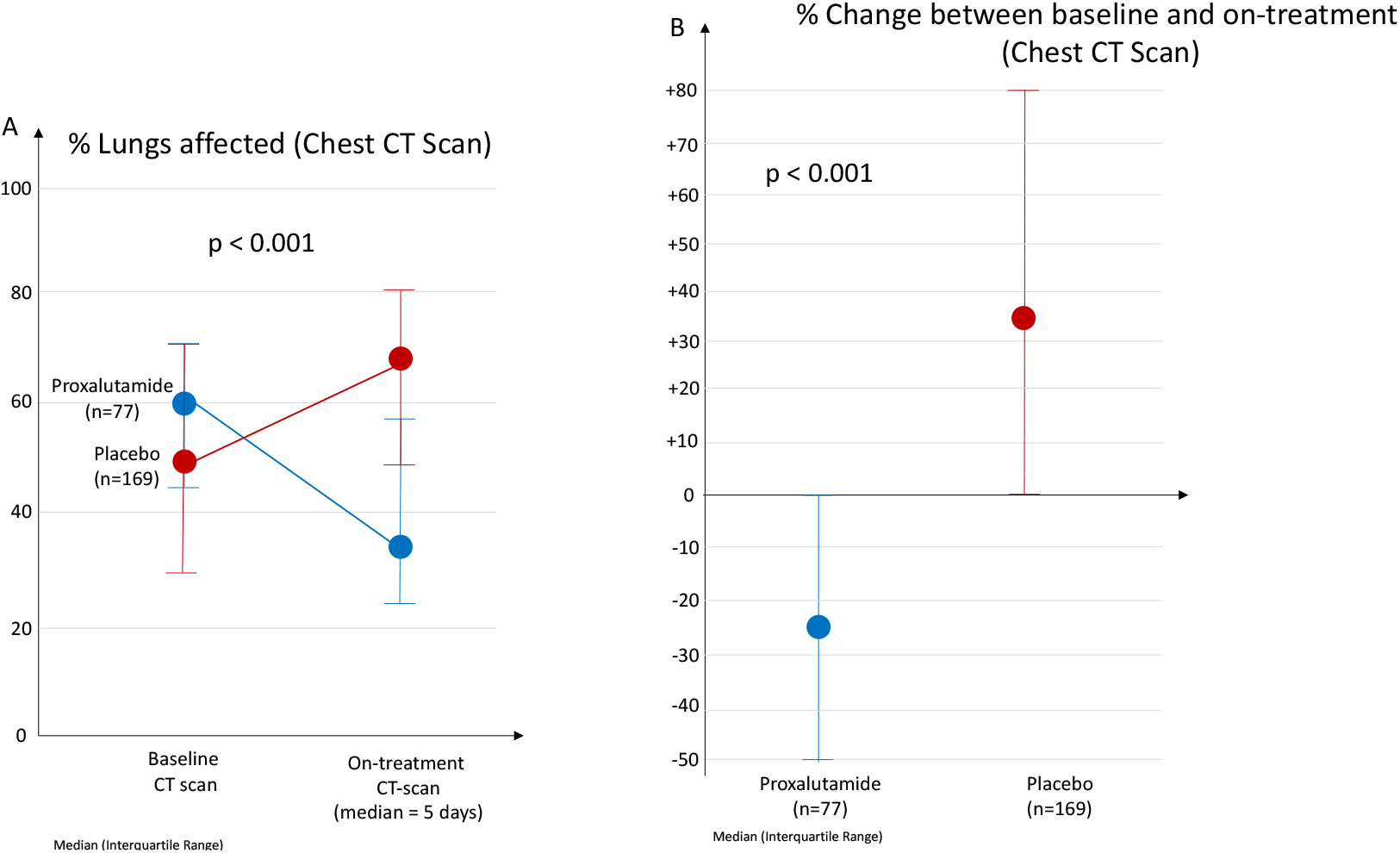
Baseline and during-treatment CT scans in proxalutamide *versus* placebo.

The percentage of lung affected (Median – IQR) in the first scan was 60% (43% - 70%) in the proxalutamide group and 50% (30% - 70%) in the placebo group (p = n/s).

In the second chest CT scan, the percentage of lungs with COVID-19 opacities (Median – IQR) was 35% (25% - 60%) in the proxalutamide group and 68% (50% - 80%) in the placebo group (p < 0.0001).

The improvement rate, *i*.*e*., the percentage of subjects that had improvement from the first to the second CT scan, was 72.3% in the proxalutamide group and 23.1% in the placebo group (p < 0.0001). The improvement rate ratio (95%CI) was 3.15 (2.32 – 4.28) with proxalutamide compared to placebo.

The change between baseline and second chest CT scans in terms of absolute percept points (pp) (Median – IQR) was -15pp (−30pp – 0pp) in the proxalutamide group and +15pp (0pp - +30pp) in the placebo group (p < 0.001).

The relative change in terms of percentage between baseline and second chest CT scans (Median – IQR) was -25% (−50% – 0%) in the proxalutamide group and +33% (0% - +80%) in the placebo group (p < 0.001).

When stratified by gender (Table 5), percentage of affected lung parenchyma second chest CT, and absolute and relative changes between baseline and second chest CT scans remained significant for all parameters for both sexes (p < 0.0001 for all). The improvement rate ratio was higher in males (5.52, 95%CI 2.83 – 7.22) than in females (2.10, 95% CI 1.39 – 3.15).

**Table 5.** Outcomes stratified by gender.

| Parameter | Gender | Overall<br>N = 246 | Proxalutamide<br>N = 77 | Placebo<br>N = 169 | p |
| --- | --- | --- | --- | --- | --- |
| Number of subjects | Female (n) | 96 | 31 | 65 |  |
|  | Male (n) | 150 | 46 | 104 |  |
| First CT scan (% of affected lungs) -<br>Median (IQR) | Female | 50 (40 – 70) | 50 (48 – 70) | 50 (30 – 70) | 0.54 |
|  | Male | 50 (40 – 70) | 60 (50 – 70) | 50 (30 – 65) | 0.02 |
| Second CT scan (% of affected lungs) -<br>Median (IQR) | Female | 60 (35 – 70) | 30 (25 – 40) | 60 (50 – 70) | < 0.0001 |
|  | Male | 60 (40 – 70) | 40 (30 – 60) | 70 (58 – 80) | < 0.0001 |
| Improvement rate - % (n) | Female | 45.8% (44) | 71.0% (22) | 32.8% (22) | < 0.0001 |
|  | Male | 34.0% (51) | 73.9% (34) | 16.3% (17) | < 0.0001 |
| Improvement rate ratio (95% CI) | Female | 2.10 (1.39 – 3.15) |  |  |  |
|  | Male | 5.52 (2.83 – 7.22) |  |  |  |
| Absolute change (pp) between first<br>and second chest CT scan - Median<br>(IQR) | Female | 0pp<br>(-20pp – +20pp) | -20pp<br>(-35pp – 0pp) | +10pp<br>(-10pp – +27pp) | < 0.0001 |
|  | Male | +10pp<br>(-15pp – +25pp) | -15pp<br>(-25pp – -1pp) | +20pp<br>(0pp – +30pp) | < 0.0001 |
| Relative change (%) in terms of<br>percentage between first and second<br>chest CT scan - Median (IQR) | Female | 0%<br>(-34% – +50%) | -33%<br>(-54% – 0%) | +20%<br>(-14% – +100%) | < 0.0001 |
|  | Male | +14%<br>(-23% – +60%) | -25%<br>(-43% – -3%) | +39%<br>(0% – +80%) | < 0.0001 |

CT = computerized tomography; IQR = interquartile range; CI = confidence interval

## Discussion

The present analysis reinforces the efficacy of proxalutamide for hospitalized patients with COVID-19. The radiological improvement provides an independent and objective evaluation of the efficacy of the drug and predicts better clinical outcomes.^12,21^ Unlike reports of clinical improvement, radiological findings analyzed by independent radiologists.

We analyzed the results utilizing the unmodified ITT, a more conservative analysis that tend to underestimate drug efficacy,^22^ in the primary analysis of the RCT. Utilizing ITT population, we observed 78% reduction in the 28-day all-cause mortality in the city of Manaus. In an on-treatment (OT) analysis, the magnitude of the results were higher (92% reduction in 28-day mortality rate).^11^. The conservative nature of the ITT analysis and the higher efficacy on treatment completers, compared to non-treatment completers, showing a “dose response-like” behavior, are also suggestive of the efficacy of proxalutamide for hospitalized COVID-19 patients, in addition to the primary findings.

To avoid selection bias, we included all participating subjects from the three hospitals of Manaus. This regional subgroup analysis maintained a balance between proxalutamide and placebo groups in terms of demographic and baseline characteristics. Samples were representative of the groups in the overall analysis. Typical limitations of a *post-hoc* analysis of a RCT appear to be absent in this study.^13^

The proportion of patients with at least two chest TC scans was significantly higher in the proxalutamide group than in the placebo group, due to increased mortality and ICU admissions in the placebo group (Table 2). Mobilization of ICU patients to CT scan became unfeasible when patients needed ICU, in particular when they were under mechanical ventilation (Table 2).

The present analysis possibly underestimates the efficacy of proxalutamide for hospitalized patients with COVID-19, for two reasons: 1. Patients that had better responses to proxalutamide did not undergo a second CT scan, since they were discharged before five days, when a second chest CT scan would be performed, as per the hospitals protocol. Consequently, a number of fast responders proxalutamide users were removed from analysis: and 2. Patients in the placebo group that had worse progression of the COVID-19 were not able to undergo a second CT scan because most of them needed ICU before five days, which precluded them from a second CT scan. In this way, patients that had better responses to usual care were probably selectively were included in the analysis. In short, this is an analysis that compared the group that responded relatively worse to proxalutamide, and therefore remained in the hospital for a longer period of time, with the group that presented a better COVID-19 disease course and did not require ICU. This hypothesis is reinforced by the fact that the median percentage of lung affected was 60.0% when all patients from the placebo group were included, and 50.0% when only those patients with at least two chest CT scans were included, while this difference did not occur in the proxalutamide group. This means that patients that were worse tended to be excluded from the placebo group. A significant improvement even under this conservative bias reinforces the potential efficacy of the drug.

The imbalance between actives and placebos in present analysis is a result of a conservative bias of the RCT: more actives were randomly designated to institutions with fewer resources in rural areas, while more placebos were randomly designated to hospital with better infra-structure. Since in-hospital mortality of COVID-19 is highly variable and largely depends on the hospital resources^23^, the use of more actives in institutions with fewer resources avoided overestimation of the drug efficacy. The analysis was conducted as Intention-To-Treat (ITT), *i*.*e*., considering patients that dropped out the study, which is another conservative bias, since the efficacy of proxalutamide in hospitalized patients largely depended on a regular and uninterrupted 14-day treatment regimen. In fact, early discontinuation of the drug is highly discouraged.

The radiologists that analyzed the chest CTs were experienced with quantifying the percentage of lungs compromised by COVID-19 as the institutions for which they worked has managed more than 20,000 cases of COVID-19,^24^ among which the vast majority underwent chest CT scans. In addition, the correlation between the quantification of COVID-19 lungs opacities by a board-certified radiologist experienced with COVID-19 chest CT-scans and artificial intelligence (AI) is strong in the majority of the cases.^25-27^ All these aspects reduce the possibility of operational-bias of the study.

The finding of radiological improvement was unexpected since the interval between the baseline and the second chest CT scan was relatively short (median of five days in both groups). We would expect that radiological changes would occur in the long-, not short-run, as per the capacity of the disease resolution, even under effective therapies.^12,20^ We hypothesize that this could be particularly true in our patient population, virtually solely infected by the Variant of Concern (VOC) P.1, arguably one of the most pathogenic SARS-CoV-2 variants described to date.^28^

In addition to the strong antiandrogen activity and to the ACE-2 antagonism, further analyses demonstrated that proxalutamide may present direct protective actions in the lungs and vessels,^7-8^ as well as anti-inflammatory effects, such as mitigation of tumor necrosis factor alpha (TNF-alpha) and nuclear factor kappa beta (NF-kB).^8^ This may explain the dramatic clinical and radiological improvements observed with proxalutamide.

Limitations of the present analysis include the fact that radiological findings are not the primary outcomes, with a subgroup of patients enrolled in three of the eight institutions that participated in the RCT. with the inherent limitations of a post-hoc analysis of a subgroup of patients of the RCT, despite the full representation of the group in the subgroup analysis and balanced characteristics between proxalutamide and placebo groups. ^11^ The present findings should be strengthened by further external analysis, in particular including an analysis using AI.

To our knowledge, this is the first RCT that demonstrated radiological improvement in response to a drug intervention in COVID-19. These findings reinforce the efficacy of proxalutamide in hospitalized COVID-19 patients.

In conclusion, proxalutamide plus usual hospitalized care demonstrated to improve hospitalized COVID-19 patients radiologically, when compared to placebo plus usual hospitalized care.

## Data Availability

Raw data is available under request to the corresponding author.

